# From Diagnosis to Demand: Obstetric Ultrasound as a Socio-Technical Practice in Rural Pakistan

**DOI:** 10.64898/2026.08.06.26359846

**Authors:** Janat Ibrahimi, Zubia Mumtaz

## Abstract

**Background:** Obstetric ultrasound is an essential tool for assessing fetal growth and wellbeing. While evidence-based recommendations advise one routine scan before 24 weeks of gestation, research suggests its use in low-and middle-income countries often extends beyond medical necessity. This study examined how ultrasound technology has become integrated into antenatal practices in rural Pakistan and how cultural, economic, and institutional factors shape its use.

**Methods:** Drawing on data from two qualitative and one mixed-method studies conducted across six rural districts of Punjab, we used a pragmatic mixed-methods approach integrating latent content analysis of interviews, focus group discussions and observations with quantitative survey data.

**Results:** Ultrasound use is common, with nearly 80 percent of women reported having at least one ultrasound and many undergoing three to six scans. For some participants, ultrasound had become synonymous with antenatal care, overtaking basic tests such as blood and urine analysis. The technology was widely perceived as both diagnostic and therapeutic, with some women believing it could cure health problems. Providers, particularly in the private sector, promoted frequent scans to meet patient expectations and financial targets, while women’s demand was driven by reassurance, perceived modernity, and son preference. This dynamic created a self-reinforcing supply–demand cycle in which clinical need played a secondary role.

**Conclusions:** In rural Pakistan, obstetric ultrasound has evolved into a central socio-technical feature of pregnancy care that extends far beyond its clinical purpose. These findings highlight the interplay of technology, culture, and market forces in shaping maternal health behaviors and underscore the need for context-sensitive policy responses that align ultrasound use with evidence-based care while engaging with the social realities that sustain its widespread adoption.

## BACKGROUND

Obstetric ultrasound is an important component of antenatal care in pregnancy management [1]. It is a useful procedure for assessing fetal growth and wellbeing, screening for fetal anomalies, locating the placenta, and assessing gestational age [2–5]. The World Health Organization recommends all mothers receive one routine ultrasound scan before 24 weeks of gestation [6].

The use of ultrasound scans in pregnancy management is an example of an introduction of a new technology in obstetric care. The World Health Organization defines technology in healthcare as “the application of organized knowledge and skills in the form of devices, medicines, vaccines, and procedures to solve a health problem and improve quality of life” [7]. A new technology must first be introduced, adopted, and seamlessly integrated into practice before it can be considered effectively utilized. Venkatesh and Bala [8] argue that potential users are more likely to adopt and continue using a technology when it aligns with their values and meets their needs.

Culture - defined as “the collective programming of the mind” [9, p. 5] through shared values, beliefs, and norms - is therefore an important determinant of whether a technology will be successfully adopted or not [9–11]. Collis [12] describes how culture influences ways in which people accept, use, and react to technology. The relationship between culture and technology is reciprocal. When a technology is created, it must first adapt into the cultural environment in order to be accepted and integrated into practice [13]. However, technology then goes onto shape the culture by influencing new social behaviours and possibilities, ultimately resulting in a social change [11,13].

Obstetric ultrasound is one example of introduction of a new health technology. As biomedically-driven pregnancy care expands in low and middle-income countries, ultrasounds have become a common part of routine antenatal care [14–19]. In Vietnam, one ultrasound scan during pregnancy is nearly universal [14]. Similarly, 98% of women in Syria [15] and 61% in India [14] reported receiving an obstetric ultrasound during their most recent pregnancy. In Uganda, a hospital-based study found that 100% of pregnant patients attending the facility received at least one ultrasound [17].

Obstetric ultrasound stands out among health-care technologies for its high level of user-engagement. Most health-care technologies require considerable technical expertise and afford little opportunity for patient involvement in their operation or interpretation. In contrast, ultrasound invites direct patient participation, particularly the latest 3-D scans that produce clear, easy-to-understand images. A large body of literature, mostly from high-income countries, has documented how these 3D ultrasound scans enable women and families to “see” their unborn baby. Mothers, their partners, and even their larger families experience this visualization positively, describing its impact in terms of increased bonding with the baby, and an enhancement of their emotional experiences of the pregnancy and impending motherhood [18–21]. So common are these activities that 3-D fetal images are now considered an integral component of mother and babyhood memorabilia [21].

However, despite the increasing use of obstetric ultrasounds, only a few studies have examined how this new technology may be impacting pregnancy management in low and middle-income countries. Emerging evidence indicates that obstetric ultrasound scans may not evoke the same benign and emotionally positive associations, such as prenatal bonding or the creation of baby memorabilia, reported in studies from high-income countries [18–21]. Instead, studies from India and Nepal show a detrimental impact on fetal sex identification and consequent abortions of female fetuses [22–24]. Some women in Vietnam, Tanzania and Rwanda have started perceiving ultrasound scans as the entirety of antenatal care [25–32]. In fact, according to Gammeltoft and Nguyen [25] and Holmlund et al [33], women in Vietnam are replacing routine antenatal care with repeated scans, some upwards of 15 scans, without receiving other elements of pregnancy care including assessment of blood pressure, body weight, hemoglobin levels or a urinalysis. These tests are crucial for the diagnosis of life-threatening conditions such as pre-eclampsia and anemia. Other studies suggest that some mothers are overestimating the diagnostic and therapeutic abilities of ultrasound technology to the point where they are not seeking or receiving appropriate care [32,33]. Tautz et. al. [33] and Firth et. al. [28] report pregnant women in Botswana and Tanzania believed that an ultrasound scan alone can treat HIV/AIDS infection. Research conducted across various countries in sub-Saharan Africa (including Rwanda, Botswana, Tanzania, and Uganda) and South-East Asia (namely Vietnam and Thailand) suggests a widespread belief that solely using ultrasound can ensure a safe pregnancy and the delivery of a “perfect baby”[26–30,32–36].

These findings are important and concerning. However, most of these studies have examined obstetric ultrasound use in isolation, without situating it within the broader social, cultural, and economic contexts that might have shaped these beliefs and practices. Specifically, obstetric ultrasound remains largely unexamined as a socio-technical phenomena, that is when a biomedical innovation becomes culturally embedded, reinterpreted and sometimes extended beyond its original clinical purpose. Accordingly, this study seeks to address the following questions: How do women interpret and use a new technology that enables visualization of the fetus in making sense of care during pregnancy? How do providers respond to women’s expectations? The present paper fills these knowledge gaps and offers a deeper insight into how uptake of a new health technology is shaped by the wider social, cultural, and economic dynamics surrounding health care.

## METHODS

This manuscript drew upon three multi-method, multi-sited studies from the senior author’s research program on maternity care in Punjab, Pakistan. The three studies explored women’s experiences of seeking maternity care, provider practices [39–40] and how health system policies and processes influence maternal health service delivery [41]. Although each study had distinct objectives within the realm of maternal health, they all, unexpectedly, produced substantive data that indicated obstetric ultrasound was being used in unanticipated ways that extended well beyond its diagnostic purposes. Collectively, data from the three studies provide an opportunity to bring multi-level, multisite qualitative and quantitative data from six districts into dialogue with one another and to examine the issue of obstetric ultrasound use through a unified analytical lens. The details of each study objectives, design, and data sources are summarized in Box.1.

***Study 1: Addressing disparities in maternal health care in Pakistan: gender, class and social exclusion* [39]**

*Objective*: To explore how gender, class and caste shape social exclusion and how these intersecting hierarchies influence women’s access to and use of maternal health care.

*Design and setting:* Critical village ethnography conducted from September 2010 to October 2014 in Chakwal district.

*Data collection:* Qualitative data were collected using multiple methods: 105 in-depth interviews, 11 focus group discussions, 134 participant observations, and longitudinal follow-up of 18 pregnant women seeking maternity services. Participants were purposively selected and included 34 young women, currently pregnant or had given birth within the past five years, 20 husbands, 27 older women, 13 older men, and 11 health-care providers offering maternal and child health services in the community. See Table 1.

*Study 2: Are community midwives addressing the inequities in access to skilled birth attendance in Punjab, Pakistan?* [40]

*Objective*: To assess whether a newly established cadre of community midwives improved rural, poor and socially excluded women’s access to maternal health care given the midwives are located in the private sector.

*Design and setting*. A mixed-methods study combining qualitative data from an institutional ethnography with quantitative data a population-based cross-sectional survey in two districts, Jhelum and Layyah. Data were collected from February 2012 to October 2016.

*Data collection:*

*1.1 Institutional ethnography*: Qualitative data were collected through 244 in-depth interviews and informal conversations and 108 observations of patient-provider interactions in district hospitals, and private practices. Participants included 68 Community Midwives, 15 Traditional Birth Attendants, 78 mothers who had given birth in the two years prior to data collection, 38 husbands, and 23 mothers and mothers-in-law.

*1.2. Population Survey*: A cross-sectional survey was conducted on a randomly drawn sample of 1,457 mothers aged 15-49 who had given birth in the two years preceding the study, 747 in Jhelum and 710 in Layyah. Using a structured questionnaire, quantitative data were collected data on socio-demographic characteristics (age, education, household income, occupation) and antenatal care indicators, including frequency and timing of ultrasound scans and routine pregnancy care laboratory investigations [40].

***Study 3: Scaling up the ‘24/7 BHU’ strategy to provide round-the-clock maternity care in Punjab, Pakistan: A theory-driven, co-produced implementation study*** [41]

*Objective*: To assess implementation of the scale-up of Pakistan’s 24/7 BHU’ initiative aimed at providing high-quality, round-the-clock skilled maternity care in first-level rural health care facilities in Punjab.

*Design and setting*. A mixed-methods study combining quantitative assessment of facility readiness, and quality of obstetric care with an Institutional Ethnography in 10 randomly selected districts in Punjab. Data were collected from January 2022 to July 2024.

*Data collection*.

An Institutional Ethnography was conducted in districts Sahiwal, Sheikhpura, Bahawalpur and Lahore. Data were collected from multiple-levels of the primary health care system, from first level care facilities, and district headquarters to ministry of health in the capital. A total 386 formal in-depth interviews and informal conversations were conducted with 49 purposively selected maternity care providers (midwives and Lady Health Visitors), 12 physicians, 78 program managers and 8 policymakers [41].

**Table 1.**
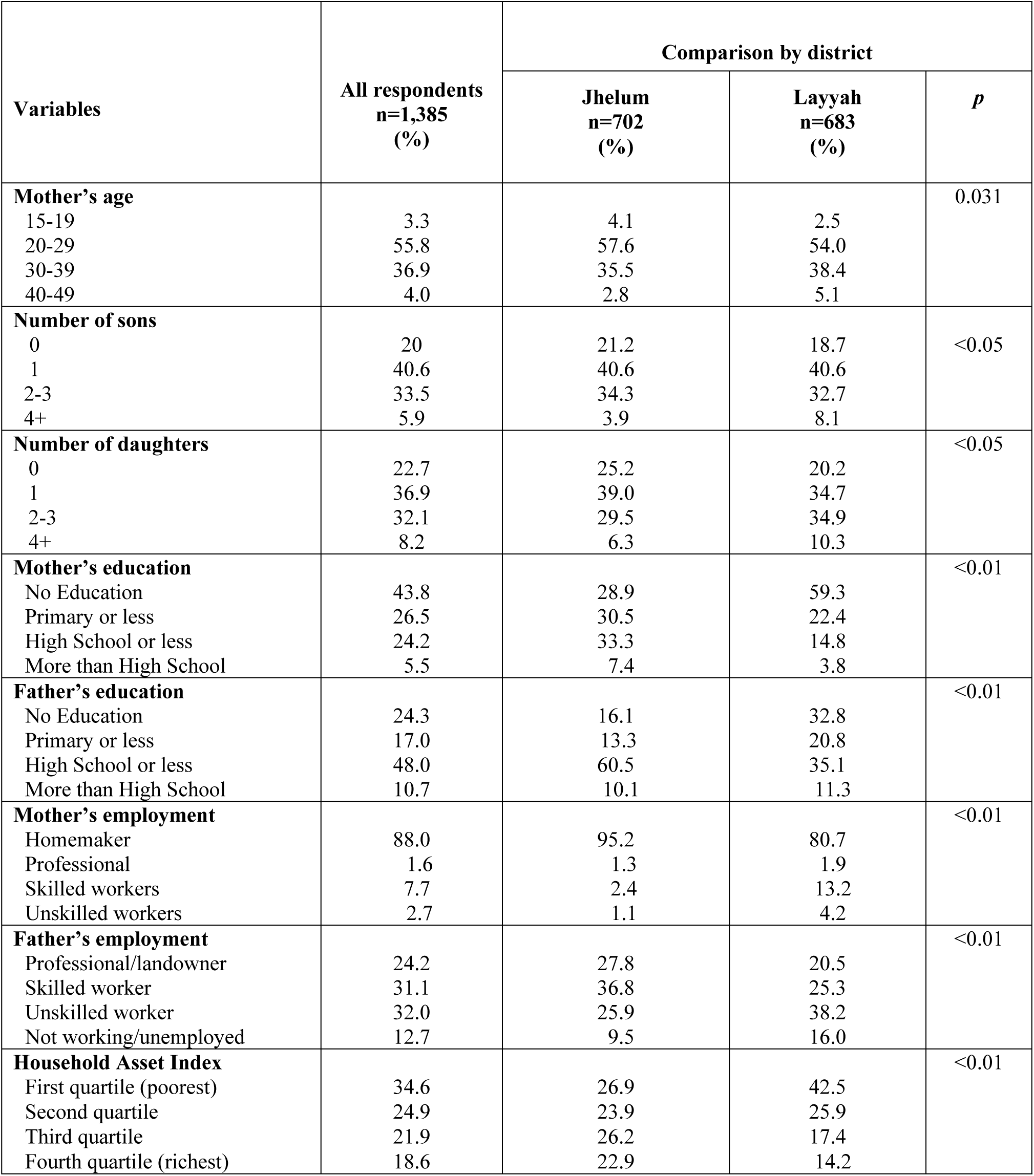
The distribution of socio-economic and antenatal care use characteristics of the survey respondents. All results are presented as percentages.

## DATA ANALYSIS

We adopted pragmatism as the guiding philosophical foundation for data analysis because it bridges realism and constructivism [42]. Pragmatism recognizes the existence of an external reality while also acknowledging the coexistence of multiple socially constructed interpretations of that reality [43]. Guided by this approach, we employed a mixed methods framework to combine qualitative and quantitative data from the three studies to provide a comprehensive understanding of how obstetric ultrasound technology functions as socio-technical tool in pregnancy management. The underlying logic of mixing qualitative and quantitative data is that they complement one another and yield a more complete analysis [44]. The qualitative data provided depth and contextual nuance in understanding the cultural meanings and lived experiences surrounding obstetric ultrasound use, whereas the quantitative data substantiated these patterns and enhanced the validity and generalizability of the findings [45].

We adopted Creswell and Plano’s [45] sequential exploratory design, treating qualitative data as the core and quantitative data as supplementary. Each data set was analyzed independently to maintain methodological integrity and consistency with its respective epistemological and ontological principles. The findings were then integrated using the weaving approach, in which both qualitative and quantitative data are presented together within a single narrative [46]. This approach yields a richer and more cohesive interpretation, thereby enhancing the validity of the findings.

### Qualitative dataset development and analysis

Qualitative data were analyzed, and reported in accordance with the Consolidated Criteria for Reporting Qualitative Research (COREQ) [47]. Given the substantive volume of data generated across the three studies, the first step was to create a focused dataset addressing obstetric ultrasound. All transcripts from the studies were systematically screened for the term ‘ultrasound’ using the search function in Microsoft Word. This process yielded 111 transcripts, which were compiled into a single dataset. This was managed and coded using Quirkos qualitative analysis software [48]. Following the principles of latent content analysis outlined by Bengtsson (2016), all transcripts were read multiple times to ensure familiarity with the data [49]. Coding proceeded in several stages: initial open coding to identify key concepts, grouping of codes into broader categories, and axial coding to explore relationships between them. Finally, through higher-level categorization, emergent themes were identified. Our latent content analytic approach allowed deeper meanings and patterns to emerge rather than imposing pre-defined categories. To maintain confidentiality, all data were anonymized, and terms “patient” and “provider” were used to denote the respective speakers.

### Quantitative dataset development and analysis

The quantitative survey data were analyzed using the statistical software package Stata 18 [50]. Since the objective of quantitative data analysis was to substantiate qualitative data patterns and not causal relationships, only descriptive univariate and bivariate analyses were done. The univariate descriptive analyses summarized the distribution of individual and household level socio-demographic, economic and antenatal care use variables including patterns of receipt of ultrasound scans and laboratory investigations (hemoglobin levels, urine tests). These summaries were presented as percentages. Bivariate analyses were then done to assess the relationship between individual-and household-level socio-demographic and economic characteristics and ultrasound use. Differences between groups were tested for statistical significance using Chi-squared tests, with a significance level set at p < 0.05.

## Ethical considerations

Ethics approval to conduct the analysis was obtained from the University of XXX No.

Pro00103670. The three original studies that collected the data received ethics approval from the University of XXX (No: Pro00019042, Pro00009329 and Pro00081045), the Pakistan National Bioethics Committee in Pakistan (No: 4-87/11/NBC/RDC/32/7 and 4-87/09/NBC-26/RDC/7881), the Provincial Bioethics Committee Punjab (No.PBC-0007) and the University of Sheffield (Reference Number 032591and 022945). It is important to state that literacy rates among women in the rural areas of Punjab, where our research was conducted, are low.

Moreover, locals commonly associate signing or thumb-printing documents with land asset transfers. Given these challenges, we received ethics clearance to obtain verbal consent from community respondents in all studies with the research team signing off the verbal consent. Written informed consent was obtained from the health care providers in the three studies. The data in all studies were anonymized and any identifiers, including names of villages or health facilities, were removed.

## RESULTS

### COMMON, POTENTIALLY EXCESSIVE USE OF OBSTETRIC ULTRASOUNDS

Both quantitative and qualitative data indicate that obstetric ultrasound use is widespread in rural Pakistan. Nearly 80% of women reported having at least one ultrasound scan during their most recent pregnancy. This widespread adoption of ultrasound is particularly striking given that 70% of the women had no formal education or only a primary education, 88% were homemakers, and nearly 60% were classified as poor or very poor (see Table 1). Notably, ultrasound use is more prevalent among younger mothers, those with higher education, and those of higher socio-economic status (see Table 2).

**Table 2:**
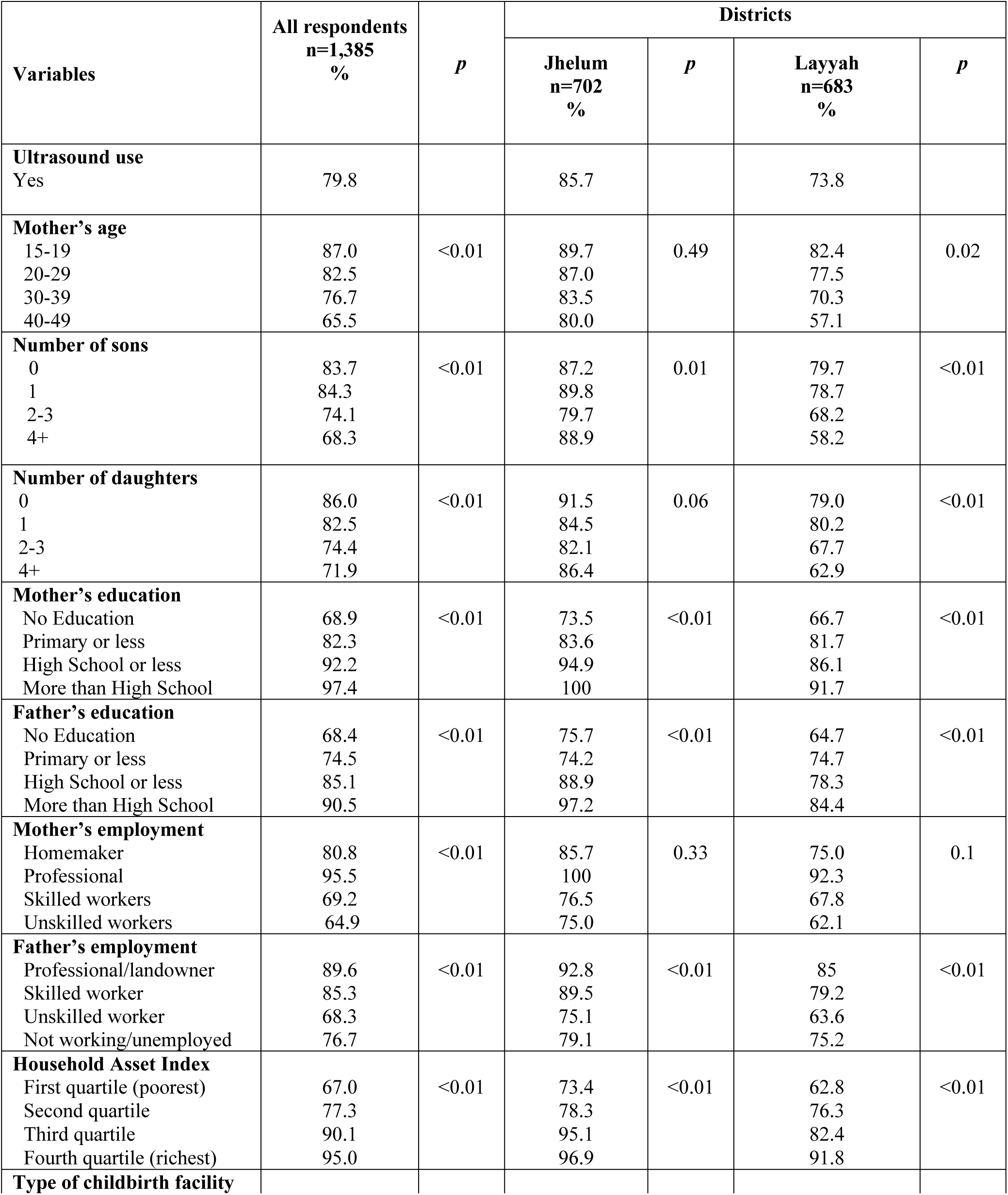
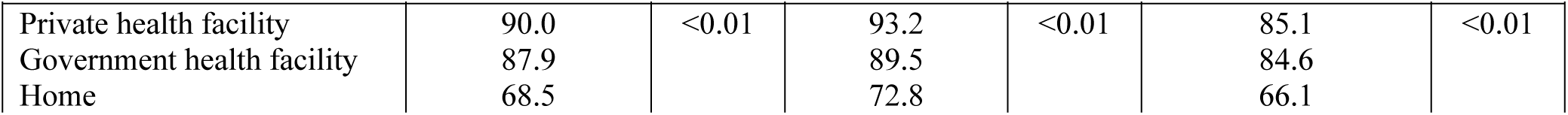
The distribution of ultrasound use by socio-economic characteristics. All results are presented as percentages.

| Variables | All respondents<br>n=1,385<br>% | *p* | Districts |  |  |  |
| --- | --- | --- | --- | --- | --- | --- |
|  |  |  | Jhelum<br>n=702<br>% | *p* | Layyah<br>n=683<br>% | *p* |
| **Ultrasound use**<br>Yes | 79.8 |  | 85.7 |  | 73.8 |  |
| **Mother's age**<br>15-19<br>20-29<br>30-39<br>40-49 | 87.0<br>82.5<br>76.7<br>65.5 | <0.01 | 89.7<br>87.0<br>83.5<br>80.0 | 0.49 | 82.4<br>77.5<br>70.3<br>57.1 | 0.02 |
| **Number of sons**<br>0<br>1<br>2-3<br>4+ | 83.7<br>84.3<br>74.1<br>68.3 | <0.01 | 87.2<br>89.8<br>79.7<br>88.9 | 0.01 | 79.7<br>78.7<br>68.2<br>58.2 | <0.01 |
| **Number of daughters**<br>0<br>1<br>2-3<br>4+ | 86.0<br>82.5<br>74.4<br>71.9 | <0.01 | 91.5<br>84.5<br>82.1<br>86.4 | 0.06 | 79.0<br>80.2<br>67.7<br>62.9 | <0.01 |
| **Mother's education**<br>No Education<br>Primary or less<br>High School or less<br>More than High School | 68.9<br>82.3<br>92.2<br>97.4 | <0.01 | 73.5<br>83.6<br>94.9<br>100 | <0.01 | 66.7<br>81.7<br>86.1<br>91.7 | <0.01 |
| **Father's education**<br>No Education<br>Primary or less<br>High School or less<br>More than High School | 68.4<br>74.5<br>85.1<br>90.5 | <0.01 | 75.7<br>74.2<br>88.9<br>97.2 | <0.01 | 64.7<br>74.7<br>78.3<br>84.4 | <0.01 |
| **Mother's employment**<br>Homemaker<br>Professional<br>Skilled workers<br>Unskilled workers | 80.8<br>95.5<br>69.2<br>64.9 | <0.01 | 85.7<br>100<br>76.5<br>75.0 | 0.33 | 75.0<br>92.3<br>67.8<br>62.1 | 0.1 |
| **Father's employment**<br>Professional/landowner<br>Skilled worker<br>Unskilled worker<br>Not working/unemployed | 89.6<br>85.3<br>68.3<br>76.7 | <0.01 | 92.8<br>89.5<br>75.1<br>79.1 | <0.01 | 85<br>79.2<br>63.6<br>75.2 | <0.01 |
| **Household Asset Index**<br>First quartile (poorest)<br>Second quartile<br>Third quartile<br>Fourth quartile (richest) | 67.0<br>77.3<br>90.1<br>95.0 | <0.01 | 73.4<br>78.3<br>95.1<br>96.9 | <0.01 | 62.8<br>76.3<br>82.4<br>91.8 | <0.01 |
| **Type of childbirth facility** |  |  |  |  |  |  |

| Variables | All respondents<br>n=1,385<br>% | p | Districts |  |  |  |
| --- | --- | --- | --- | --- | --- | --- |
|  |  |  | Jhelum<br>n=702<br>% | p | Layyah<br>n=683<br>% | p |
| Private health facility | 90.0 | <0.01 | 93.2 | <0.01 | 85.1 | <0.01 |
| Government health facility | 87.9 |  | 89.5 |  | 84.6 |  |
| Home | 68.5 |  | 72.8 |  | 66.1 |  |

Qualitative findings also revealed that ultrasound has become a routine and expected component of pregnancy management, with both women and healthcare providers frequently referring to it as a standard part of antenatal care. The data further suggest that the frequency of obstetric ultrasounds often exceeds formal guidelines. Ultrasound use in Pakistan is guided by WHO recommendations of one routine scan before 24 weeks of pregnancy. However, most participants reported undergoing three to six scans during an otherwise uncomplicated pregnancy.

> *“Interviewer: How many times did you get your ultrasound test done? Woman 1: Three to four times”” (Mother, aged 25-29 years, Bahawalpur)*.

In fact, ultrasound scans have become so commonplace that women reported receiving one at nearly every antenatal care (ANC) visit, most commonly during monthly appointments. Health care providers confirmed this pattern, explaining that the monthly ultrasounds were a more effective means of monitoring pregnancy than traditional abdominal palpation. Many also framed this practice as a response to women’s expectations, noting that women wanted ultrasounds as part of their antenatal examination. The providers expressed concern that if they did not comply with these demands, they would risk “losing” patients to other providers who were willing to perform the expected scans. For private providers, the loss of patients was a loss of revenue, and for government health facilities, an inability to meet their antenatal care targets.

> *“But people think that antenatal care means that you will have to get your ultrasound done every time you come for checkups” (Midwife)*.

Moreover, our analysis of the data by a physician (the senior author) suggests the majority of these ultrasound scans could be deemed medically unnecessary. All the 18 mothers followed longitudinally had normal pregnancies, yet they received an ultrasound scan at every antenatal care visit, which varied from 4 to 6 visits. One woman reported undergoing four scans due to “low blood pressure.” Another mother had multiple scans because her husband had a blood group B Rh-negative while hers was blood group A Rh-positive. This combination does not pose a risk for Rh incompatibility, as Rh incompatibility occurs when an Rh-negative mother carries an Rh-positive fetus. Another woman justified the frequent scans saying that she had “katchra” in her uterus, which literally translates into “garbage” in the uterus. However, some cases of frequent scans were legitimate as in the case of a woman who stated her baby had “growth” issues.

So ingrained is the idea of a scan at every antenatal visit that mother often determined a provider’s quality of care simply in terms of whether they received an ultrasound scan. For example, many mothers described feeling satisfied with their provider’s quality of care solely because they conducted an ultrasound scan during every antenatal examination. Conversely, the absence of a scan left mother feeling deprived of comprehensive or high-quality care.

> *“I am not totally satisfied with the quality of service here because they didn’t do an ultrasound scan. I am not feeling good about it.” (Mother, aged 25-30, Chakwal)*

The providers mused on the reasons underlying the quick adoption of ultrasound technology in an otherwise conservative region that has a history of resisting many public health endeavours. One view was that women understood the technology as a signifier of modernity and progress, that they were using new, more ‘scientific’ and advanced forms of pregnancy care. Ultrasound machines were first introduced in major tertiary care hospitals in big cities, which are viewed as sites of modern medical care.

> *“The problem is these ultrasound machines were first introduced in large teaching hospitals in cities, not in small private clinics in rural areas. And foreigners have introduced these machines in our country and our people got influenced by this” (Physician, 30-35 years, Sahiwal)*

Other providers mused that ultrasound scans signalled a higher social status. In rural areas ultrasound scans were first introduced in and even now are usually only available in private health facilities which primarily served wealthier individuals who could afford the associated costs. Earlier on, the test was quite expensive and it was mostly the well-off who could afford such care.

> *“Actually, the ultrasound machines were used in private hospitals, not in government BHUs. The upper class used to go to these private hospitals” (Midwife, 35-40 years, Bahawalpur)*.

All this has now set a stage where women experience peer pressure to seek ultrasound scans. The peer pressure can be so strong that poor mothers borrow money for the test.

> *“Women experience peer pressure…Why haven’t you had your ultrasound yet? You’re five to six months into your pregnancy; it should have been done by now. Is someone preventing you from getting it done? Is it your mother-in-law or other family members?…This social pressure drives many people to seek ultrasounds”* (Physician, 25-30 years, Sahiwal)

## UNINTENDED EFFECTS OF OBSTETRIC ULTRASOUNDs

### Ultrasound scans as the entirety of antenatal care

An unintended consequence of the widespread use of obstetric ultrasound scans is that mothers are starting to understand an ultrasound scan as constituting the entirety of antenatal care. When asked what antenatal care they received, women invariably answered “*an ultrasound…what else*”. We observed women seeking care and asking for ‘*machine lagao’* (apply the machine). The equation between antenatal care and ultrasound was so widespread that mothers used the term ‘check-up’ to refer to both the antenatal visit and the ultrasound scan.

Healthcare providers corroborated these findings, commenting that “*people think an ultrasound is part of their checkup*”. Several midwives expressed concerns about their clients’ tendency to regard ultrasound scans as the primary aspect of antenatal care, and that they simultaneously devaluated other elements of antenatal care, including essential procedures such as blood pressure measurement, urinalysis for proteinuria, or blood tests for hemoglobin levels. When mothers were probed if they had blood or urine tests, the majority replied in the negative, arguing they did not need these tests as they were healthy.

These qualitative findings are substantiated by the quantitative data, which show that while 80% of the mothers reported receiving an obstetric ultrasound scan, a lower proportion, specifically 49.6%, reported having received a blood test for anemia, and 48.6%, reported having undergone a urine test for proteinuria. These data become more pronounced in the less-developed district of Layyah, where 73% of mother received ultrasound scans, far surpassing the percentages for urine and blood tests (24% and 27% respectively). This suggests that nearly 50% of mother in Layyah only received an obstetric ultrasound scan as the only test during their last pregnancy. See Table 3.

**Table 3.** The distribution of elements of antenatal care received as reported by survey respondents. All results are presented as percentages.

| Variables | All respondents<br>n=1,385<br>(%) | Comparison by district |  |  |
| --- | --- | --- | --- | --- |
|  |  | Jhelum<br>n=702<br>(%) | Layyah<br>n=683<br>(%) | *p* |
| **Sought one or more ANC visit** | 95.6 | 97.2 | 93.9 | 0.03 |
| **ANC provider** |  |  |  |  |
| Doctor | 36.5 | 38.2 | 34.6 | <0.01 |
| Trained Midwives | 47.9 | 53.3 | 42.2 | <0.01 |
| Traditional Birth Attendant | 8.7 | 4.5 | 13.3 |  |
| Other unskilled birth attendants | 6.9 | 4.1 | 9.9 |  |
| **Elements of ANC received** |  |  |  |  |
| Ultrasound scan | 79.7 | 85.6 | 73.5 | <0.01 |
| Body weight measured | 49.3 | 66.4 | 31.4 | <0.01 |
| Urinalysis | 48.7 | 72.3 | 24.1 | <0.01 |
| Blood testing (for haemoglobin) | 49.6 | 71.1 | 27.0 | <0.01 |
| Tetanus injection | 90.2 | 92.5 | 87.8 | <0.01 |
| Iron supplements prescribed | 83.6 | 88.1 | 78.8 | <0.01 |
| Dietary advice | 80.1 | 87.0 | 72.8 | <0.01 |
| Information of danger signs of pregnancy | 41.4 | 52.4 | 29.9 | <0.01 |
| Developed a plan in the event of an emergency | 36.0 | 43.9 | 27.8 | <0.01 |
| Discussion around importance of breastfeeding | 70.3 | 77.8 | 62.5 | <0.01 |
| Family planning | 45.5 | 50.1 | 40.2 | <0.00 |

### Potentially misplaced trust in ultrasound technology

One factor driving the frequent and often unwarranted use of ultrasound scans was the profound confidence mothers had in the technology’s capacity to detect health issues in both the mother and the baby. Women sought the scans for reassurance that the baby was well, but also as a comprehensive diagnostic tool for any health issues they were experiencing. Providers described examples of pregnant mother requesting an obstetric ultrasound to assess “a gas-like pain in the stomach”, their “level of blood”, and to “examine the position of my bones*”*. In fact, the data suggests some mothers believed an ultrasound was even therapeutic, with requests for a scan to treat urinary tract infections, and even hasten labour. There were instances of mother calling for the machine to be “applied” in order to remedy complications during labour.

> *“Whenever, I feel unwell, I apply the machine (referring to getting an ultrasound scan) and take the medicine” (Mother, 25-30, Bahawalpur)*

So strong was this trust in the powers of ultrasound technology, that mothers expected everything reported by the test to be precise, including the date of expected birth and determination of fetal sex. Providers shared the challenges that came with this level of trust and how any discrepancies between ultrasound predictions and actual outcomes resulted in significant disappointment. If a baby’s delivery date did not exactly match the one stated in their ultrasound report, mother and their families were, at most, annoyed or inconvenienced. However, fetal sex reports had much more serious implications, both for providers and mothers. Some hospital staff reported experiencing aggression and physical threats of violence from families when a daughter was delivered instead of an expected, ultrasound-scan-determined son. For mothers, a wrong fetal sex disclosure – in other words, the birth of a daughter instead of a son – potentially had devastating effects on her emotional and physical well-being. Although rarely vocalized, but clearly insinuated, the key reason many mothers sought ultrasound scans was to find out the fetal sex. To be more precise, it was to confirm that the fetus was male. Pakistan is a context characterised by strong son preference. The importance of sons is woven in the society’s social, economic and gender order. Not having a son can destabilize a woman’s position within the household and shape her relationships with her extended family. Mothers in our study who did not have a son felt incomplete, that they have not fulfilled their duty as a woman or a wife. They were at risk of divorce and abandonment.

> *“When the doctor told the family that a girl instead of the expected boy, the father shouted at her that her mother had been giving birth to daughters, and that she would also do the same. He then kicked her out of his house and told her and not come back to his house until she was able to give birth to a boy.” (Mother, aged 30-35, Chakwal)*

## SUPPLY-DEMAND CYCLE OF UNNECESSARY OBSTETRIC ULTRASOUND SCANS

The qualitative data suggest both providers and mothers/family members are active participants in driving the use of possibly unnecessary obstetrics ultrasounds scans. Below, we explore the two broader societal forces that have contributed to the reinforcing cycle of supply and demand for unnecessary obstetric ultrasound scans.

### Private sector marketing ultrasound scans for profit

The data show that the perception of ultrasound scans as a crucial element of antenatal care and the strong trust mothers’ place in the technology has been deliberately cultivated by medically trained providers. These providers have leveraged on their position and authority as a doctor – with all its attendant clout as an expert – to create a lucrative market for ultrasound technology. They took advantage of the social and cultural value of son-preference described above to actively shape a demand for ultrasounds, thereby creating a profitable market niche. Despite their rationalization that their frequent use of ultrasound scans was a response to the patients’ demands and that it was driven by peer pressure, we empirically observed providers actively recommending and conducting an ultrasound scan as a standard component of a routine antenatal visit. We observed providers prioritize an ultrasound scan over other elements of antenatal care such as measuring body weight or blood pressure. This was also noticed by the one patient who pointed out that *“No one does blood or urine tests; these doctors only do ultrasound.” (Mother, aged 30-35, Chakwal)*.

The private sector providers did most of this marketing because upon making the investment in an ultrasound machine, the only way of recouping that investment is through an increase in the volume of scans performed. Some providers went further and aggressively advertised their ultrasound scanning services, even offering discounts such as a scan for only PKR 100 (USD $1.00) every last Friday of the month. This was a significant discount from the usual rates of PKR 500-700 (UDS 5.00 – 7.00). Some midwives working in government health facilities invested in personal mobile ultrasound machines, which they brought to the government facility and charged the patients, often under the table. Some even got the local mosque to advertise their ultrasound services as a community service, but at a cost. Many had created networks in which a group of providers, including the Traditional Birth Attendants and other unskilled providers send referrals and accepted ultrasound reports only from each other. Those who did not have an ultrasound scanning machine would refer to specific members of these networks for a commission. If a mother sought care from a provider outside the network, she would be required to get another ultrasound from a member of the informal network, increasing her costs.

> *“They charge 200 rupees per patient for an ultrasound scan. The hospital keeps 150 and gives the referring provider 50 rupees.” (Observation notes, Layyah)*

The private sector’s success in the promotion of ultrasound technology is validated by the quantitative evidence, which shows the high rate of ultrasound use at nearly 80%. Notably, this marketing has been effective across a broad spectrum of socioeconomic groups. Both the rich and poor mothers report its use, albeit at different rates of 95% and 67% respectively. There is variation by district, with Jhelum mothers reporting higher rates of ultrasound. This aligns with overall greater development and wealth of the district and ability of mothers to pay for private sector maternity care and ultrasound scans.

### Patients and providers are mutually reinforcing the use of unnecessary ultrasound scans

Our data suggest pregnant mothers are active participants in driving the unnecessary scans. Mothers chose clinics for antenatal care based on the availability of ultrasound machine and services in the facility. Providers described many instances of losing clients to competitors solely because they lacked ultrasound machines.

> *“The Community Midwives (a part of a public-private initiative) were demanding ultrasound machines from the government. They said that patients don’t seek their care because they do not have ultrasound machines.” (Observation notes, Chakwal)*

Mother’s demand for ultrasound scans was primarily driven by strong son-preference described above. They went to great lengths to ensure they gave birth to a son. The mothers sought repeated ultrasound scans not just to reassure fetal health, but actually to either confirm that the sex of the fetus was indeed male or in the event of a female diagnosis, to rule out an error. Mothers would continue seeking scans from different providers – also called “doctor shopping” among providers – if the fetal sex report did not match their preferred sex. They specially sought care from providers who were rumoured to deliver only sons.

> *“She said she expects to have a son this time because she is going to Dr. X for an ultrasound scan.” (Research Team Observations notes, Chakwal)*.

Our study providers drew upon this deep cultural need for a son to expand their business. While disclosing the sex of the fetus is not legally prohibited, it is generally discouraged because of the ethical concerns surrounding fetal sex selection. Nonetheless, our observations showed that providers were willing to disclose fetal sex when requested by patients. The providers were even more forthcoming in revealing the sex of the fetus if it was male, while showing hesitancy and suggesting additional scans for female fetuses. According to these providers, they risk losing clients if they report a female fetus. Further exploiting the cultural biases and mother’s vulnerabilities around son-preference for financial gain, the providers charged hefty fees for revealing fetal sex because the benefits of knowing fetal sex heavily outweighed any high costs for often desperate mother. Certain providers were even reported to be charging more for a scan reporting a male fetus.

> *“It happened a couple of times that a patient insisted on knowing the baby’s sex. It was her first baby. She fell silent when I told her it was a girl. I never saw her again”. (Provider, Chakwal)*

## DISCUSSION

This study demonstrates how obstetric ultrasound in rural Pakistan has transformed from a purely diagnostic instrument into a normalized feature of antenatal care, one that simultaneously embodies cultural values, reinforces gender hierarchies, and generates economic profit within the healthcare market. While the original objective of ultrasound scans was monitoring maternal and fetal well-being, the technology has evolved into a socio-technical practice symbolizing modernity, social status and provisions of “high-quality” care. It has blurred the boundaries between an ostensibly objective medical technology and cultural meanings ascribed to it, transforming its purpose to fit local expectations and needs in ways that have, at times, rendered its original clinical purpose secondary.

The integration of qualitative and quantitative evidence highlights the scale of this transformation. Nearly 80% of mothers surveyed reported undergoing at least one ultrasound during their most recent pregnancy, a considerably higher proportion than those who reported having blood or urine tests. Qualitative data revealed that many women received three to six scans for uncomplicated pregnancies, reflecting a pattern of routine, and often unnecessary, use.

Similar trends have been reported from many countries in the Global South. Women in Vietnam reported receiving an average of 6.6 scans in 8.3 antenatal visits [14]. Another survey from reported women receiving more than 15 scans without any other forms of pregnancy management [51]. Similar findings were reported from Syria, and Iran, with women receiving an average of 5.5 scans and 5.9 scans respectively during their most recent pregnancy [7, 8].

The perception of ultrasound as the entirety of antenatal care, and trust in the technology’s ability to offer thorough and complete care during pregnancy, even potentially being curative, was another prominent finding in this research. Women described the scan as capable of detecting “anything wrong” with themselves or the fetus, and some believed it could treat ailments or hasten labor. Similar faith in ultrasound technology has been documented in diverse contexts spanning Asia and Africa, including Vietnam, Tanzania, Rwanda and Botswana [25, 27, 28, 29]. Midwives in Hanoi reported that some pregnant women were replacing routine antenatal care with repeated scans. Some of these women were even described to ignore any other antenatal clinical examinations in favour of an ultrasound scan [29]. Similarly, women in Botswana, described ultrasound as a “magical device that can see everything” [34]. The Pakistani experience thus aligns with evidence from other culturally varied contexts, suggesting that beliefs and values surrounding reproductive technologies that extend beyond their original clinical purpose are more the norm than the exception.

The trust women place in ultrasound technology also potentially reflects an epistemological shift from relational, observation-based care to visual-technological authority, where the image replaces clinical judgment. The phenomenon shows how medical technologies can redefine knowledge hierarchies by elevating the screen over the stethoscope and the machine over the provider, while reshaping how women interpret health and risk. The research also demonstrates how medical technology can be mobilized to serve deeply rooted social expectations. It highlights the intersection of gender unequal values and technology, demonstrating ways in which women and society have appropriated ultrasound technology to reproduce and reinforce son preference. The technology enabled the fulfillment of desires that were previously beyond human reach, transforming what was fate regarding the sex of the fetus into a seemingly controllable outcome.

The data suggests a prominent driver of the socio-technical practices around ultrasounds is the economic structure of Pakistan’s health care system, a mixed model in which the majority market-driven private-sector services funded through out-of-pocket payments co-exists with publicly financed government services [52]. The public sector operates a spatially distributed three-tier health care system, but inefficiencies in delivery of care fueled a neoliberal donor-driven expansion of the private sector over the past 40 years [53]. Consequently, the private sector now accounts for over 70% of health expenditure [52, 54]. It provides the majority of maternity care, delivering nearly 70% of antenatal services and servicing over two-thirds of facility births [55]. This sector has strategically capitalized on both the perceived indispensability of obstetric ultrasound and on its symbolic power [51,56,57]. Private providers have made ultrasound scanning a center-piece of antenatal visits, promoting it as a sign of quality while marginalizing basic examinations such as blood or urine testing. This commercialization of technology is not unusual for it mirrors similar trends in other health services in countries such as Vietnam, where profit motives—not medical necessity—determine frequency of use [51,56,57].

The findings of this research point to the need for thoughtful and contextually grounded policy responses that recognize how ultrasound technology is deeply intertwined with social life and health practices. Such policies should acknowledge that these changes represent an entrenched socio-technical and cultural shift rather than misuse of technology. They should avoid imposing externally conceived notions of ‘appropriate biomedical use’ and instead engage with how women, providers and communities value and integrate the technology into their understanding of maternity care. Nevertheless, there is room for policy to prioritize strengthening clinical training and quality assurance to align ultrasound use with evidence-based practices, while expanding public education that emphasizes the full spectrum of antenatal care.

The study has some limitations. The first is that it was an analysis of data that was collected for other objectives. The findings presented in this paper surfaced unexpectedly during an inductive, open-ended and exploratory analysis of the qualitative data. The quantitative survey had only asked if the respondents received an ultrasound, not their frequency. We were, therefore, unable to measure the frequency of ultrasound scans. The survey data also did not collect information on the private-sector location of the antenatal care providers, which emerged as a key determinant of excessive and unnecessary scans. Given these limitations, we used Creswell and Plano’s [45] sequential exploratory design, treating qualitative data as the core and quantitative data as supplementary. We recommend further research to explore the levels and determinants of qualitative findings. The second limitation of our study is that the primary qualitative data were collected in six districts, and quantitative data from two districts. While the latter two districts represent the spectrum of overall socio-economic development, we cannot extrapolate the data to the whole of Pakistan, or even the province of Punjab. The third limitation is that women might have under-reported blood or urine tests which might not be as memorable as an ultrasound. While possible, our just concluded, not-yet-published research on quality of maternity care in rural Pakistan shows that not all women receive the recommended blood and urine tests. The fourth limitation is that we did not investigate clinical situations that might have necessitated more than the recommended number of ultrasounds. The data collection took place in a village ethnography setting, characterized by low literacy rates among women and a health system that did not maintain clinical records. The final limitation is that the qualitative data were translated from Urdu to English with the risks of not capturing the full meaning of the data. This risk was mitigated by the PI of all three studies, who is fluent in both Urdu and English. The PI read through the Urdu transcriptions and English translations of a subsample of data twice.

## CONCLUSION

This study reveals that obstetric ultrasound use in rural Pakistan has expanded far beyond its original clinical purpose, evolving into a central socio-technical component of antenatal care. It illustrates how a biomedical technology, initially introduced to enhance pregnancy monitoring, has become culturally embedded in ways that reshape both mothers’ expectations and provider practices. These patterns underscore the complex interplay between technology, culture, and market forces in shaping reproductive health practices.

## Data Availability

The data are available at https://era.library.ualberta.ca/items/027e1ac8-4629-48e2-8501-795610921da7

